# Hospital costs associated with mechanical left ventricular mechanical unloading devices during VA ECMO for adult cardiogenic shock

**DOI:** 10.1101/2025.03.31.25325000

**Authors:** Maxwell A. Hockstein, Joshua J. Horns, Thomas Hanff, Iosef Taleb, Craig Selzman, Stavros Drakos, Richard Nelson, Joseph E. Tonna

**Author notes:** <u>Corresponding Author:</u> Joseph E Tonna, MD, MS, 30 N Mario Capecchi Drive, Level 4 South, Salt Lake City, UT 84112,; Twitter: @JoeTonnaMD. <u>Authorship:</u> Study design: JJH, JT; Study conduct: JJH, JT; Data acquisition and analysis: JJH, JT; Drafting the manuscript: MH, JJH, JT; all authors revised the article for important intellectual content, had approved the final manuscript for publication. JT had full access to the study data and takes responsibility for the data integrity, accuracy, and integrity of the submission as a whole.

## Abstract

**Importance:** No works to-date have described the financial burden and behaviors of left ventricular mechanical unloading (LVMU) for patients on veno-arterial extracorporeal membrane oxygenation (VA ECMO). Given the uptrending use of VA ECMO, describing its associated cost is essential for its continued uptake.

**Objective:** We describe the inpatient costs of patients who were managed with ECMO for cardiogenic shock (CS) with and without LVMU.

**Design, Setting and Participants:** We conducted a retrospective cohort study of adult (age ≥18 years) patients who received ECMO at some point during their hospital stay and were non-post operative patients (e.g. medical CS) using the IBM MarketScan database. Data were extracted from 1/1/2008-12/31/2021.

**Exposures:** The exposure of interest was the additional use of LVMU (intra aortic balloon pump, peripherally inserted left ventricular assist device [pVAD], or “other”) added to ECMO. Costs were calculated daily, and modeled according to the daily status of ECMO, LVMU, ECMO+LVMU, or no device.

**Main Outcomes and Measures:** Patient characteristic, including age, sex, comorbidities quantified using the Charlson Comorbidity Index (CCI), etiology of heart failure (acute myocardial infarction [AMI] vs chronic heart failure [CHF]), hospital and intensive care unit (ICU) length of stay (LOS) and total inpatient costs were described using descriptive statics between groups. The outcomes of interest were total inpatient costs. Secondary outcomes included in-hospital mortality, and hospital and intensive care unit (ICU) length of stays (LOS). We stratified patients by receipt of LVMU, and used propensity score matching from patient level characteristics to balance the use of LVMU between groups. Cost outcomes were modeled using mixed effects linear regression clustered on matched groups and reporting incident rate ratios (IRR). LOS and mortality outcomes were modeled using Poisson (IRR) and logistic (adjusted odds ratio [aOR]) regression, respectively, conditional on matched groups.

**Results:** Enrolled patients (n=1,596) were 56 years old (interquartile range [IQR] 47 to 62), had an ICU LOS of 9 (3 to 19) days, and a hospital length of stay of 18 (7 to 35) days, which were not different between groups. Patients who received LVMU had a higher CCI (*p<*0.001), and were more likely to have a primary CS etiology of AMI (54% vs 39%; *p*<0.001) but not CHF (66% vs 62%; *p*<0.08). The median total inpatient cost of ECMO alone was $320,269 vs $390,508 (ECMO+LVMU [*p*<0.001]). In adjusted analysis, compared to patients without ECMO or LVMU, the daily incurred costs for patients on ECMO alone were three times higher (cost ratio = 3.0, p<0.001), 2.6 times higher for patients on LVMU alone (cost ratio = 2.6, p<0.001), and 4.2 times higher for people on both ECMO and LVMU (cost ratio = 4.2, p<0.001). Patients with ECMO+LVMU had a longer hospital LOS (IRR 1.059; *p*<0.001) compared to ECMO alone, but a similar hospital ICU LOS (IRR 0.98; *p=*0.08). Patients who received LVMU had significantly lower mortality than those who only received ECMO (HR = 0.62, p=0.006).

**Conclusions and Relevance:** CS patients managed with VA ECMO + LVMU had significantly increased cost, significantly longer LOS, and significantly decreased mortality compared to ECMO alone. Understanding the impact of LVMU on the cost of an ECMO course will aid appropriate resource allocation.

## INTRODUCTION

Treatment success for the use of extracorporeal membrane oxygenation (ECMO) has traditionally been defined as survival to ECMO explant or to hospital discharge. This definition of success is justified, as avoidance of mortality is generally agreed to be a foundational goal of life, not just of medicine. Equally as important as examining ECMO’s contributions to outcomes, is quantifying its economics as relevant to uptake and sustainability. Consequently, as the use of ECMO has increased over time, with well over 30,000 annual runs across age groups worldwide, attention has increasingly been directed at the finances of ECMO treatment.^1^

A successful ECMO course requires the coalescence of multiple service lines, professionals, medications, processes, and technologies for a single patient’s survival. As a result, patients on ECMO incur significant costs associated with their hospitalization. The financial impact of ECMO for respiratory failure was examined in the landmark CESAR trial in 2009 and during extracorporeal cardiopulmonary resuscitation (ECPR). ECMO has been generally identified as financially favorable, but few studies have examined the costs associated with veno-arterial (VA) ECMO treatment for cardiogenic shock (CS).^2–7^

Patients on ECMO for CS are at particular risk of incurring high costs due to the prevalent use of concomitant devices to mechanically unload the left ventricle, which has been consistently associated with improved survival during ECMO, and for which three randomized clinical trials are underway (NCT06336655, NCT05913622, NCT05577195).^8^ Despite the observed survival association, no study to-date has examined the financial burden of left ventricular mechanical unloading (LVMU) during ECMO.

To address this gap, we performed a retrospective analysis of an insurance claims database to examine the hospital costs, length of stay and mortality associated with the addition of LVMU devices to adult patients on VA ECMO for CS.

## METHODS

### Data Source and Study Population

We conducted a retrospective cohort study using the Merative MarketScan database. The MarketScan database contains de-identified commercial claims encounter data presently accessible by institutional subscription; MarketScan utilizes data from patients living in the United States with commercial insurance or Medicare parts C and D. The data includes dates, diagnoses, procedures, and costs associated with inpatient healthcare encounters. We extracted data from 1/1/2008-12/31/2021, including patients if they were ≥18 years and received ECMO during at least part of their hospitalization. If the same patient had two separate but contiguous admission records that both involved ECMO, the records were combined. Patients were deemed to have undergone LVMU if they received a peripherally inserted left ventricular assist device (pVAD), an intra-aortic balloon pump (IABP), or another LVMU device during their ECMO course. (**Supplemental Table 1**)

Patients were excluded from the analysis cohort if they had incomplete demographic information, three or more separate ECMO admission records, if they underwent a cardiac operative procedure during the admission, or if their ECMO duration was less than 24 hours. This later exclusion was to exclude patients who had ECMO only for peri-procedural support (coronary angiography, percutaneous coronary interventions, etc). The presence, timing, and duration of concomitant LVMU was recorded. Patients were analyzed on a daily basis and categorized as having one of the following states: 1) ECMO alone, 2) ECMO+LVMU, 3) IABP or pVAD alone, 4) no device.

### Outcomes

The primary outcome was total accrued inpatient cost. Secondary outcomes included hospital length of stay (LOS) and intensive care unit (ICU) length of stay (ILOS). Mortality was analyzed using data from years during which the database collected mortality (2008-2015).

### Statistical Analysis

Variables were summarized using median and interquartile range (IQR) for continuous variables and counts and percentages (%) for categorical variables. We performed univariate tests of significance on all demographic and hospital-stay variables stratified by whether patients received LVMU during the course of their ECMO. We used chi-squared tests or Mann-Whitney U tests as appropriate.

For analysis of the primary outcome, patients were stratified by receipt of LVMU. To balance other factors against the receipt of LVMU, we utilized propensity score (PS) matching. Propensity scores of receiving LVMU were constructed using sex, age, CCI, acute diagnosis of acute myocardial infarction (AMI), and or chronic heart failure (CHF) exacerbation as cofactors. Patients were compared with replacement at a 5:1 ratio (no LVMU: LVMU) with a caliper of 20% of the standard deviation (SD = 0.12). All covariates included in the PS model were well-balanced post-matching. Standardized mean differences before and after matching are shown in **<u>Supplemental Figure 1</u>**.

For the mortality models we ran a Cox proportional hazards model conditional on the propensity score (PS) matching. T_0_ was set at time of ECMO initiation and patients were censored when they were discharged. For the LOS outcomes, we performed Poisson regression conditional on matched PS groups. For cost outcomes we utilized mixed-effect linear regressions clustered on matched PS groups and patient ID. We examined daily costs and categorized each 24 hour day as one in which the patient received an additional LVMU or not. Only patients with known ECMO and LVMU durations were included. Costs were adjusted for inflation and log-transformed prior to modeling. Missing data are reported in **Supplemental Table 2**. Results were reported according to STROBE guidelines. All statistical work was performed with R version 4.4.0.^9^ P values <0.05 were considered statistically significant.

## RESULTS

### Study Population

The database query identified 5,063 patients who received VA ECMO from 1/1/2011-12/31/2021, with 1,291 patients subsequently excluded. A participant flow diagram is seen in **Figure 1** and cohort characteristics are seen in **Table 1**. After propensity matching, the final analytic cohort consisted of 1,596 patients. Among patients, 31.2% were female. In unadjusted analysis, age, hospital length of stay, and ICU LOS did not significantly differ between the 434 (27.19%) patients who underwent LVMU and the 1162 (72.81%) patients without LVMU.

**Table 1.** Cohort demographics.

|  | ECMO alone | ECMO+LVMU | Full Cohort | p-value |
| --- | --- | --- | --- | --- |
| **Total** | **1162 (72.8%)** | **434 (27.2%)** | **1596 (100%)** |  |
| Age | 56 (46 to 62) | 57 (49 to 62) 0 | 56 (47 to 62) | **0.123** |
| LOS | 17 (6.25 to 35) | 19.5 (9 to 34.75) | 18 (7 to 35) | **0.118** |
| ICU LOS | 9 (3 to 20) | 10 (3 to 19) | 9 (3 to 19) | **0.62** |
| Total Payment (US dollars) | 320,269 (149,807 to 636,243) | 390,508 (201,463 to 774,996) | 343,467 (165,383 to 670,680) | **<0.001** |
| **Sex** |  |  |  | **0.148** |
| Male | 787 (67.7%) | 311 (71.7%) | 1098 (68.8%) |  |
| Female | 375 (32.3%) | 123 (28.3%) | 498 (31.2%) |  |
| **CCI** |  |  |  | **0.001** |
| 0 | 131 (11.3%) | 21 (4.8%) | 152 (9.5%) |  |
| 1 | 261 (22.5%) | 85 (19.6%) | 346 (21.7%) |  |
| 2 | 241 (20.7%) | 104 (24%) | 345 (21.6%) |  |
| 3+ | 529 (45.5%) | 224 (51.6%) | 753 (47.2%) |  |
| **Comorbidities** |  |  |  |  |
| AMI | 456 (39.2%) | 235 (54.2%) | 691 (43.3%) | **0.001** |
| CHF | 714 (61.5%) | 288 (66.4%) | 1002 (62.8%) | **0.08** |

**Abbreviations:** AMI-acute myocardial infarction; CCI-charlson comorbidity index; CHF-chronic heart failure; ICU-intensive care unit; LOS-length of stay; US-United States
*Note: P values denote the result of the Kruskal-Wallis or Chi-Squared test as appropriate.*

**Figure 1.**
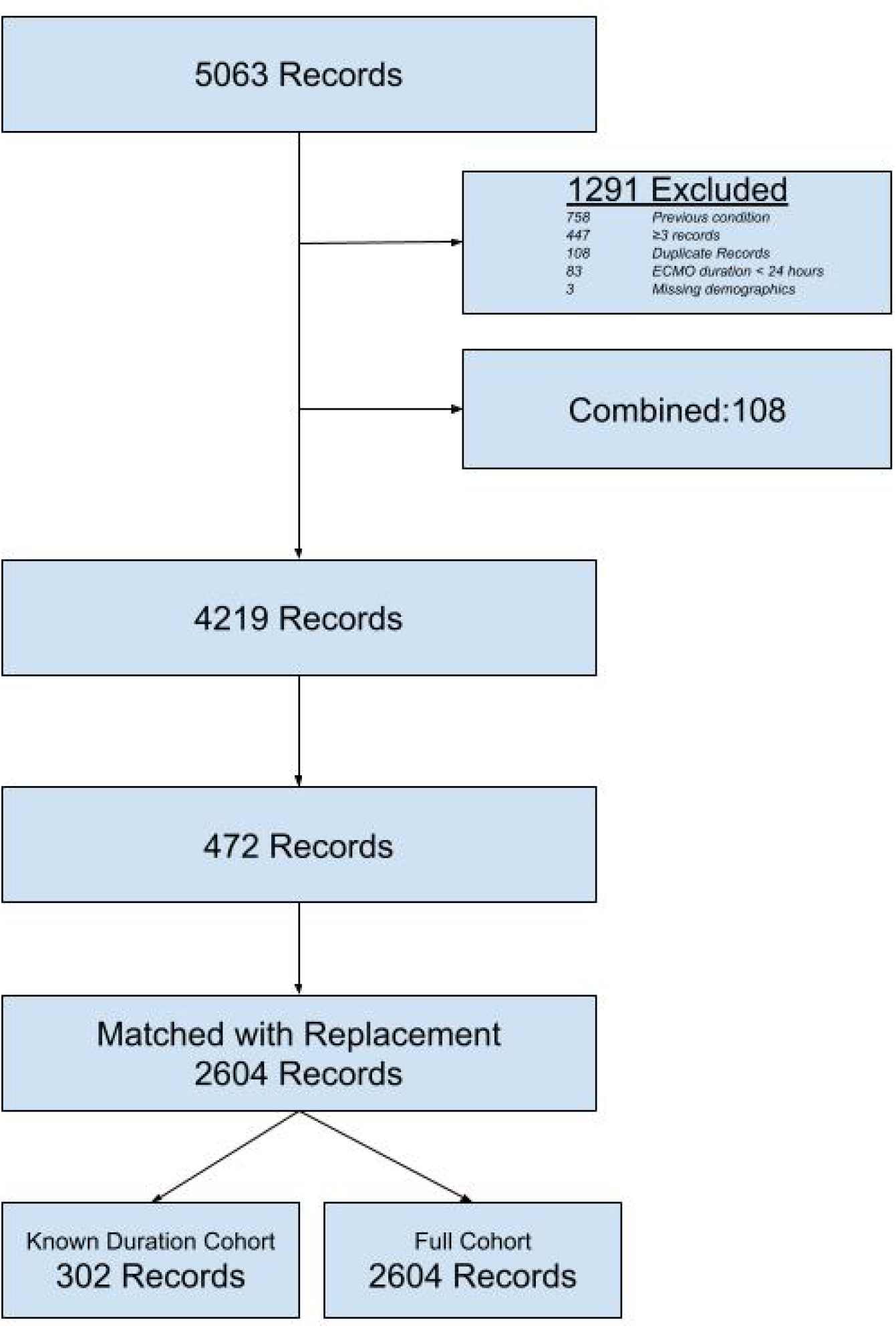
Cohort flow diagram.

### Total Cost

The median total cost for the entire admission in patients who only received ECMO was $320,269 and the median total inpatient cost of patients who received both ECMO and LVMU was $390,508 (a 21.9% increase) **(Table 1)**. When analyzing the cost of each day separately, there was a cost hierarchy based on the devices; the use of IABP/pVAD alone had the lowest cost (IRR 2.6) compared to days in which neither ECMO nor LVMU were used. ECMO alone had the next highest cost (IRR 3.02), followed by ECMO+LVMU as the highest cost (IRR 4.2) as seen in **Table 2** and **Figure 2A**.

**Table 2.** Association of LVMU on total daily cost.

|  | IRR | 95% CI | p-value |
| --- | --- | --- | --- |
| Intercept | 798.6 | (712.4, 895.2) | <0.001 |
| ECMO | 3.016 | (2.805, 3.242) | <0.001 |
| ECMO+ LVMU | 4.200 | (3.302, 5.341) | <0.001 |
| IABP or pVAD alone | 2.610 | (1.846, 3.688) | <0.001 |

**Abbreviations:** CI-confidence interval; CCI-Charlson Comorbidity Index; ECMO-extracorporeal membrane oxygenation; IRR-incident rate ratio; LVMU-left ventricular mechanical unloading

**Figure 2.**
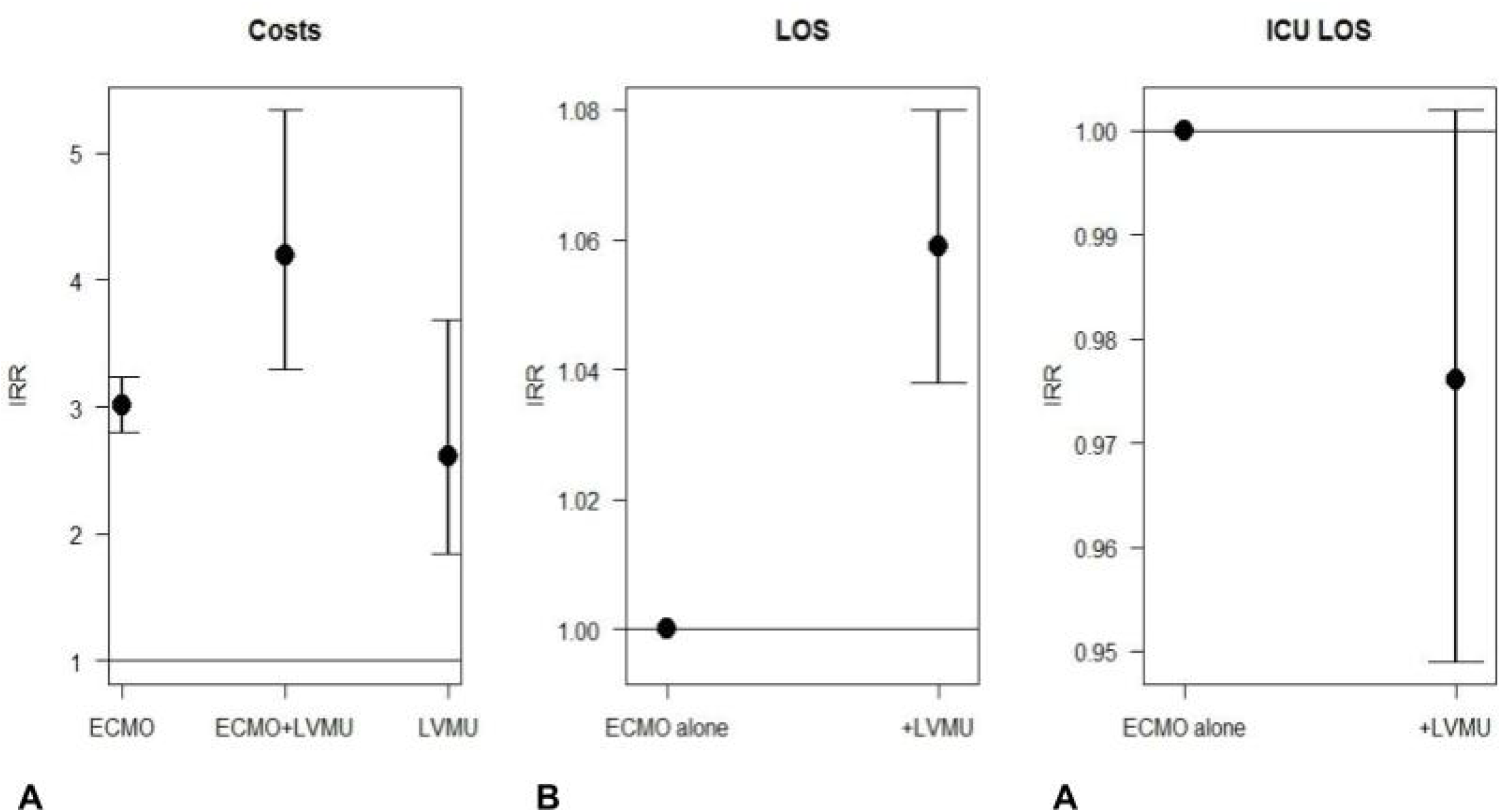
Panel A demonstrates incident risk ratios of costs between support modalities. Panels B and C demonstrate the incident risk ratios of total hospital and ICU lengths of stays, respectively.

### ICU and Hospital Length of Stay

Total hospital LOS was significantly longer in patients who underwent ECMO+LVMU compared to those with ECMO alone (ECMO+LVMU IRR 1.06 [1.04, 1.08; p<0.001]). ICU LOS was non-significantly shorter among patients who received ECMO+LVMU compared to those with ECMO alone (ECMO+LVMU IRR 0.98 [0.95, 1.0; *p*=0.076]) (**Table 3, Figure 2B and 2C**).

**Table 3.** Association of LVMU on total hospital and ICU length of stay.

| Length of Stay |  |  |  |
| --- | --- | --- | --- |
|  | IRR | 95% CI | p-value |
| ECMO | *reference* |  |  |
| ECMO+LVMU | 1.059 | (1.038,1.080) | <0.001 |
| Intensive Care Unit Length of Stay |  |  |  |
|  | IRR | 95% CI | p-value |
| ECMO | *reference* |  |  |
| ECMO+LVMU | 0.976 | (0.949,1.002) | 0.076 |

**Abbreviations:** aOR-adjusted odds ratio; CCI-charlson comorbidity index; CI-confidence interval; ECMO-extracorporeal membrane oxygenation; ICU-intensive care unit; IRR-incident risk ratio; LVMU-left ventricular mechanical unloading

### Impact of LV Unloading on Mortality

The analysis examining the association between LVMU during ECMO and mortality identified 2744 records, which after propensity matching resulted in 929 patients. Patients who received LVMU had significantly reduced mortality (HR = 0.62 [0.44-0.87], p=0.006) compared to patients who only received ECMO (**Figure 3, Supplemental Table 3**).

**Figure 3.**
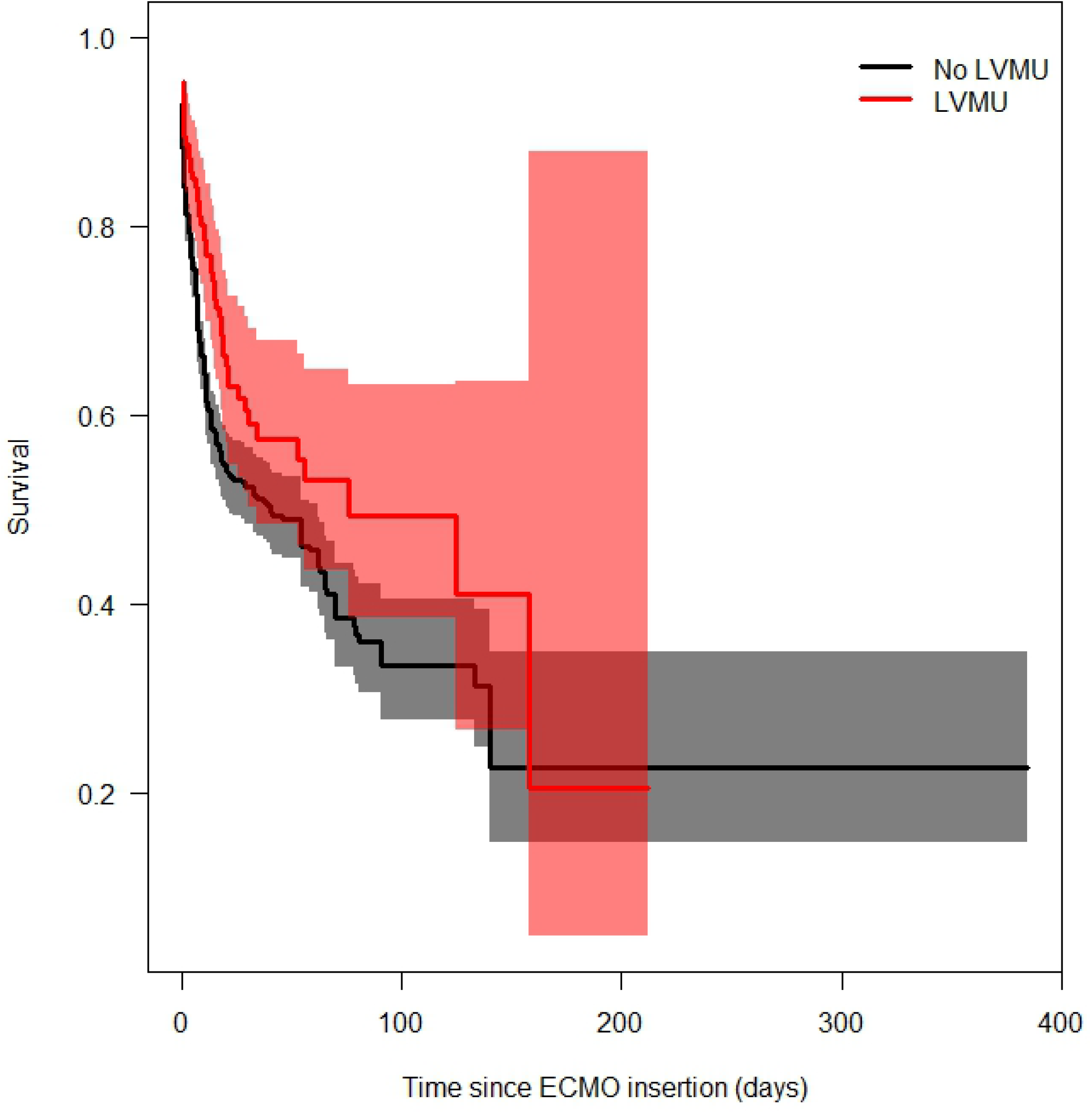
Kaplan Meier survival estimates for patients who received ECMO + LVMU versus ECMO alone

## DISCUSSION

Our analysis demonstrates that the receipt of concomitant LVMU for patients who underwent VA ECMO for non-post operative CS resulted in increased costs, increased durations of hospitalization, improved mortality and similar ICU LOS compared to patients managed with VA ECMO alone. Patient days with any device (ECMO, IABP/pVAD, or ECMO+LVMU) had increased daily costs compared to days with no device, but the cost of a day with ECMO was similar to the cost of day with IABP or pVAD alone.

Although reported costs are often inconsistent across institutions or regions, hospitalizations for CS are broadly understood to be costly and prevalent.^10^ CS hospitalizations requiring ECMO are significantly more so.^11^ Despite this, limited studies have examined cost or cost effectiveness for treatment with VA ECMO for CS or comparisons of cost using different approaches. Indeed, prospective observational studies are ongoing to evaluate the cost effectiveness comparison of Impella® vs VA ECMO for CS.^1^ Among published studies, the use of VA ECMO for CS while costly, is believed to be cost effective.^2,3^

Despite the observed survival association and findings of cost effectiveness from VA ECMO for CS, other studies have reinforced the findings of high expense, even suggesting a lack of cost effectiveness for select populations, such as bridge to transplant.^4–6^ The use of Impella® alone has mixed data on cost, especially compared to the use of the less expensive IABP.^7–11^ Both devices are expensive. Historically, IABP alone has not demonstrated a survival advanced when used alone as mechanical circulatory support, though the recently published DanGer Shock trial did show a benefit to Impella® alone for infarct related shock.^12,13^ Likewise VA ECMO, which a shown benefit in cardiac arrest resuscitation, has not yet shown clear mortality benefit in CS.^14–17^

Against this backdrop, with ongoing clinical trials to evaluate the effect of LVMU added to VA ECMO (NCT06336655, NCT05913622, NCT05577195), the costs and clinical utilization associated with adding an LVMU device to ECMO are unknown. Our findings help fill this gap by showing in a large national administrative insurance claims dataset that patient day costs are incrementally related to the use of mechanical circulatory support devices, increasing from none, to IABP/pVAD alone, to ECMO alone, to ECMO+LVMU. The use of LVMU added to ECMO significantly increases costs of ECMO alone, but is also associated with a significant improvement in mortality.

These findings are logical, based on our understanding of the benefit to decompressing/unloading the left ventricle, especially in the setting of AMI, with or without VA ECMO.^18,19^ Ventricular unloading has demonstrated benefit in a chronic heart failure model, and our findings add further data that mechanical ventricular unloading during VA ECMO has a benefit to myocardial recovery.^20,21^ The non-significant reduction in ICU LOS experienced by the group who underwent unloading is likely multifactorial. In addition to plausible myocardial recovery and thus faster inotropic wean, patients who received LV unloading may have experienced less pulmonary edema and therefore had a diminished need for mechanical ventilation. Alternatively, patients who received LV unloading had shorter ECMO courses than their controls.

We acknowledge several limitations. The MarketScan database is an incomplete repository of healthcare claims, not including patients with Medicare parts A or B, TRICARE, or Indian Health Service. Given the older age of medicare patients, and the known use of ECMO in older patients, it is likely that many patients who received ECMO are not included.^12^ Secondly, given the incomplete capture of other medical claims outside of MarketScan, patients who changed insurance off of a plan captured by MarketScan would necessarily have their data omitted, leading to underestimation of healthcare costs. If this change in insurance was imbalanced between groups, it would introduce bias. Further, as patients were only included if they had placement and removal CPT codes for all devices, as it is known that not all removals are coded as robustly as placement, we likely had multiple patients excluded who did in fact undergo unloading during ECMO but were excluded. We acknowledge that LV venting practices evolved over the study period and consequently rates of each LV venting strategy were not constant. Finally, as the MarketShare database does not explicitly collect baseline variables such as cardiac function, or hemodynamics, and/or contraindications to LV venting, we were unable to adjudicate the diagnosis of cardiogenic shock or balance of severity of shock, which is known to correlate with mortality.^22,23^

## CONCLUSIONS

Our investigation is first to describe the daily and accumulated inpatient healthcare costs associated with the use of concomitant LV unloading for patients on VA ECMO using a large healthcare claims database. Our findings show that LV unloading added to VA ECMO was associated with increased inpatient costs, longer LOS and improved mortality compared to ECMO alone.

## Data Availability

Data is available upon reasonable request.

## Abbreviations

VA ECMO: venoarterial extracorporeal membrane oxygenation

---

**Supplemental Table 1.** Exclusionary criteria.

| Criteria | Number |
| --- | --- |
| Post operative status | 758 (14.97%) |
| Missing demographics | 3 (0.06%) |
| >3 ECMO Runs | 447 (8.83%) |
| ECMO duration less than 24 hours | 83 (1.64%) |

**Abbreviations:** ECMO-extracorporeal membrane oxygenation

**Supplemental Table 2.** Missing data.

Missing data
| Variable | Full Cohort n=1596 |
| --- | --- |
| Age | 0 (0%) |
| LOS duration | 0 (0%) |
| ICU LOS duration | 0 (0%) |
| Total Payment | 365 (22.8%) |

**Abbreviations:** ICU-intensive care unit; LOS-length of stay

**Supplemental Table 3.**
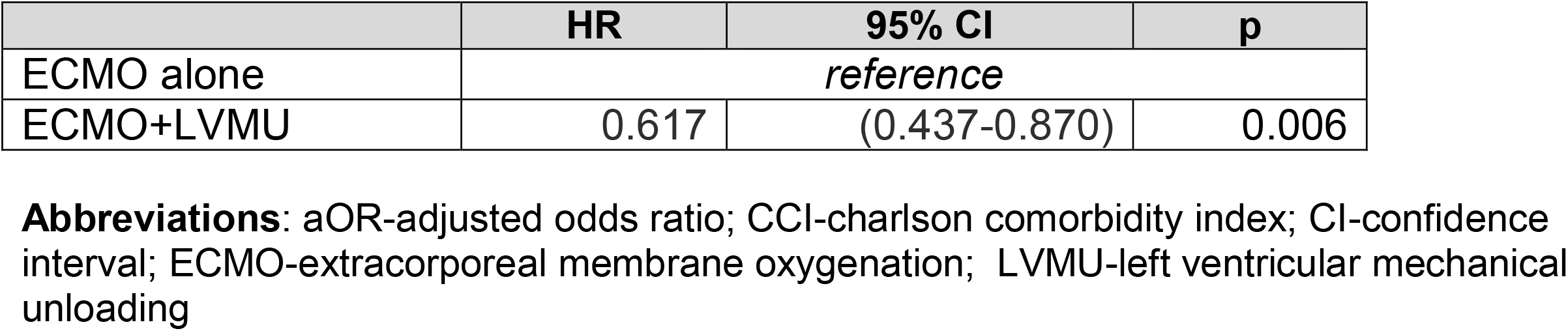
Adjusted Mortality from Cox Proportional Hazard Model with ECMO+LVMU.

**Supplemental Figure 1** – Standardized mean differences before and after matching

## Notes

<u>Conflicts of interest and source of funding:</u> Dr. Tonna is supported by R01HL168510 from the National Institutes of Health (NIH)/National Heart Lung and Blood Institute (NHLBI). None of the funding sources were involved in the design or conduct of the study, collection, management, analysis or interpretation of the data, or preparation, review or approval of the manuscript. No conflicts of interest reported.

### Competing Interest Statement

The authors have declared no competing interest.

### Funding Statement

No external funding.

